# Evaluating Resting State EEG Biomarkers Across Psychosis Biotypes: Stability and HD-tDCS Modulation

**DOI:** 10.64898/2026.02.23.26346924

**Authors:** Rebekah L Trotti, Isaac W Doss, David A Parker, Nicolas Raymond, Kodiak Sauer, Godfrey D Pearlson, Elliot S Gershon, Sarah K Keedy, Scot K Hill, Carol A Tamminga, Jennifer McDowell, Paulo Lizano, Matcheri S Keshavan, Brett A Clementz

## Abstract

**Objective:** We examined the clinical utility of resting state electroencephalography (rsEEG) by evaluating its temporal stability, discriminant validity for B-SNIP psychosis Biotypes, and suitability as a treatment target for brain stimulation.

**Methods:** We collected 5 minutes of eyes-open rsEEG from 1401 participants with psychosis and 750 healthy persons. A subset of participants was re-tested after 6 months and 12 months (N=109). In a pilot target engagement study (n=5) we collected rsEEG before and after 2 high-definition transcranial direct current stimulation (HD-tDCS) interventions targeting the left dorsolateral prefrontal cortex (dlPFC) and temporoparietal junction (TPJ). Data were reduced with principal component analyses to delta/theta, alpha, beta, and gamma frequency bands, and compared between groups and timepoints.

**Results:** rsEEG frequency bands displayed good-to-excellent stability and significantly distinguished psychosis Biotypes with large effect sizes. Compared to healthy, Biotype-1 had low activity (average ES=-.58), Biotype-2 had high activity (ES=1.07), and Biotype-3 had slightly elevated activity (ES=.33). There were no rsEEG differences between DSM psychosis groups. After anodal dlPFC stimulation, alpha and gamma power slightly increased while positive symptoms and verbal fluency improved. After cathodal TPJ stimulation, delta/theta power slightly increased while psychoticism and digit sequencing improved.

**Conclusions:** Resting state brain activity is a trait-like marker that differentiates B-SNIP psychosis Biotypes, suggesting differing underlying neurophysiology. The pilot intervention supports the feasibility of targeting this underlying neurophysiology with HD-tDCS. Integrating rsEEG in diagnostic procedures and stratified intervention selection may be beneficial for psychosis patients.

## Introduction

The brain’s spontaneous activity at rest is a product of its structural integrity, network dynamics, and neural noise [1]. This intrinsic activity affects the signal-to-noise ratio of the brain, where the neural response to incoming stimuli (signal) must be appropriately in-balance with the ongoing background neural activity (noise) to promote veridical perception, environmental appraisal, and cognitive and behavioral responses. Using non-invasive EEG recordings during an eyes-open or eyes-closed resting state (rsEEG) gives researchers and clinicians a simple, portable, and low-cost biomarker of intrinsic neural activity, which could have clinical utility for psychiatry.

Impaired signal-to-noise ratio may be a feature of psychosis, with consequences for sensory fidelity and cognitive performance [2,3]. Difficulty parsing biological and clinical heterogeneity in psychiatry limits the utility of rsEEG as a psychosis biomarker. A review of rsEEG publications across several different psychiatric conditions including schizophrenia found that measures have very little diagnostic specificity, small effect sizes, and weak symptom correlations [4]. Similarly, a previous Bipolar-Schizophrenia Network for Intermediate Phenotypes (B-SNIP) publication [5] evaluated pseudo-resting state EEG extracted from 10-second rest intervals between auditory stimuli in a large sample of participants with schizophrenia, schizoaffective disorder, and bipolar disorder. This study found no significant differences from healthy persons or between DSM diagnostic groups. Still, understanding variation in rsEEG appears highly relevant to psychiatry, as other studies suggest that it is associated with genetic risk for psychosis [6] and could be affected by psychiatric drugs like clozapine [7].

Intrinsic neural activity may be an important biomarker for persons with psychosis. An important characteristic of treatment-relevant biomarkers is temporal stability in the absence of interventions aided at modifying their magnitudes. In healthy adults, conventional rsEEG metrics show good-to-excellent test-retest reliability, with highest reliability for scalp-level oscillations in the alpha band [8]. Likewise, Clementz et al. [9] showed a repeatability of .87 for ongoing high frequency activity, a related although not equivalent measure of background brain signal [10]. Despite evidence for rsEEG stability in healthy persons and on related measures, subtle psychosis subgroup variations could affect reliability in an otherwise stable psychosis population. We aim to examine the test-retest reliability of rsEEG in a clinically stable population without treatment changes to determine the degree of rsEEG stability over time.

The approach of biological subtype identification in psychosis may be particularly helpful for developing rsEEG as a useful biomarker. Prior studies indicate that B-SNIP psychosis Biotypes have robust differences in pseudo-rsEEG extracted from rests between stimuli [5,9–11], which captures the same latent construct as pure resting state (intrinsic neural activity).

Results showed that intrinsic neural noise is abnormally low in Biotype-1 and abnormally high in Biotype-2, differences that are canceled if groups are determined by DSM psychosis diagnoses. While these characteristics were identified in pseudo-rsEEG, pure resting state recordings like those used for neurological testing are more easily collected and clinically translatable, as they do not require a stimulus, temporally precise equipment, or significant processing and storage resources. A primary goal of this paper is to replicate intrinsic EEG differences between psychosis Biotypes during a short, task-free state of rest.

Intrinsic measures also can be longitudinally evaluated during an intervention to examine their value as treatment targets. Though stability in the absence of intervention is critical to qualify a biomarker as a valid target for intervention, this feature does not mean they must be immutable. Other biomarkers used in medicine (e.g., HbA1c) show high test-retest reliability yet remain modifiable. One promising technology for targeting electrophysiology is high-definition transcranial electrical stimulation (HD-tES), a noninvasive technique wherein electrodes placed on the scalp deliver low-intensity electrical currents to a region of cortex. With direct current, anodal stimulation increases and cathodal stimulation decreases the underlying membrane potential of the targeted region. Though research is still in early stages, tES seeks to target and normalize electrophysiological biomarkers in clinical populations. We examined rsEEG’s value as a treatment target for HD-tES by evaluating it before and after 2 common tES interventions for psychosis [12,13]: anodal stimulation of the dorsolateral prefrontal cortex (dlPFC) and cathodal stimulation of the temporoparietal junction (TPJ). These treatments target dlPFC hypoactivity related to cognitive impairments [14–16] and TPJ hyperactivity related to social and positive symptoms [17,18].

This work aims to evaluate the translational potential of rsEEG by 1) demonstrating test-retest reliability of rsEEG in a stable psychosis population, 2) replicating distinctive patterns of intrinsic activity in B-SNIP psychosis Biotypes, and 3) examining rsEEG changes induced by 2 HD-tES interventions. We hypothesize that rsEEG frequency measures will 1) display high test-retest reliability in a sample with no change in treatments, 2) differentiate psychosis Biotypes from one another and healthy persons (Biotype-1 < Healthy/Biotype-3 < Biotype-2), and 3) be enhanced in the frontal cortex by anodal HD-tES to the dlPFC and attenuated in the posterior parietal cortex by cathodal HD-tES to the TPJ. Support for the first two hypotheses would indicate that rsEEG reveals trait-like biomarkers of deficient and excessive background brain activity underlying two types of psychosis pathology. The third hypothesis would provide preliminary proof-of-concept evidence that rsEEG could be an effective target for novel treatment development and test the feasibility of such an intervention.

## Method

This study encompasses 3 components: the longitudinal stability sample, the cross-sectional sample, and the HD-tES target sample. The longitudinal and HD-tES components re-recruited participants from the cross-sectional sample and used identical inclusion/exclusion criteria, clinical assessments, EEG procedures, and EEG processing methods. Prior cross-sectional B-SNIP papers have examined intrinsic neural activity derived from event-related EEG [5,10,11], but this is the first to examine Biotype differences in a pure resting state.

### Participants

The longitudinal stability sample included B-SNIP2 participants recruited from 5 academic and medical institutions across the United States. 80 with psychosis and 29 healthy persons were evaluated at baseline, 6 months, and 12 months. The cross-sectional sample included 3 B-SNIP cohorts: B-SNIP1, PARDIP (Psychosis and Affective Research Domains and Intermediate Phenotypes), and B-SNIP2, collected at 7 institutions in the United States. 1401 participants with psychotic disorders and 750 healthy comparisons were assessed (Table 1, Tables S1-2). Race, ethnicity, and sex were ascertained by participant report. The tES target sample included 5 B-SNIP2 participants in Boston, MA only, evaluated before and after each intervention with an open-label crossover design. The Institutional Review Boards at each institution approved the projects and participants provided written informed consent prior to study enrollment.

**Table 1.** Cross Sectional Demographics.

| **Characteristic** | **Healthy**<br>**N = 750** | **B1<sup>1</sup>**<br>**N = 476** | **B2<sup>1</sup>**<br>**N = 447** | **B3<sup>1</sup>**<br>**N = 478** | **SZ<sup>1</sup>**<br>**N = 583** | **SAD<sup>1</sup>**<br>**N = 439** | **BD w/P<sup>1</sup>**<br>**N = 379** |
| --- | --- | --- | --- | --- | --- | --- | --- |
| **Mean Age (SD)** | 35 (12) | 37 (12) | 39 (11) | 36 (12) | 37 (12) | 39 (12) | 36 (12) |
| **Sex (%)** |  |  |  |  |  |  |  |
| Male | 43% | 60% | 44% | 51% | 64% | 45% | 42% |
| Female | 57% | 40% | 56% | 49% | 36% | 55% | 58% |
| **Ethnicity (%)** |  |  |  |  |  |  |  |
| Not Hispanic | 88% | 84% | 86% | 87% | 88% | 83% | 85% |
| Hispanic | 12% | 16% | 14% | 13% | 12% | 17% | 15% |
| **Race (%)** |  |  |  |  |  |  |  |
| Black/African American | 31% | 49% | 41% | 23% | 49% | 37% | 20% |
| American Indian | 0.3% | 0.8% | 0.2% | 0.2% | 0.3% | 0.5% | 0.5% |
| Asian | 8.2% | 2.8% | 2.5% | 3.6% | 4.1% | 2.5% | 1.6% |
| White/Caucasian | 53% | 39% | 46% | 64% | 38% | 47% | 71% |
| Native Hawaiian/<br>Pacific Islander | 0.1% | 0% | 0% | 0% | 5.2% | 9.0% | 4.5% |
| Multiracial | 4.3% | 5.7% | 7.0% | 5.9% | 0% | 0% | 0% |
| Other | 2.8% | 3.2% | 3.6% | 3.6% | 3.3% | 3.9% | 3.2% |
| **Mean Global Functioning (SD)** | 85 (7) | 52 (12) | 51 (12) | 57 (13) | 50 (12) | 53 (12) | 59 (13) |
| **Mean SES of Individual (SD)<sup>3</sup>** | 35 (14) | 50 (14) | 49 (14) | 42 (15) | 50 (14) | 48 (14) | 41 (14) |
| **Mean SES of Family (SD)<sup>3</sup>** | 38 (15) | 44 (16) | 44 (16) | 37 (16) | 43 (16) | 43 (16) | 37 (16) |

<sup>1</sup>B=Biotype; SZ=Schizophrenia, SAD=Schizoaffective Disorder, BD w/P=Bipolar Disorder with Psychotic Features
<sup>3</sup>Hollingshead Index of Socioeconomic Status Score (higher scores = lower SES)

Participants were administered the Structured Clinical Interview for DSM Diagnosis (SCID-IV-TR) [19]. Clinically stable outpatients were diagnosed with schizophrenia, schizoaffective disorder, or bipolar I disorder with a history of psychotic features. Healthy persons had no history of psychosis, mania, or recurrent depressive episodes and had no family history of psychosis or bipolar disorder. Exclusion criteria for all participants included current illicit drug use (established by urine drug screening), alcohol or substance abuse within 1 month or dependence within 3 months of the study, and any major neurological, cognitive, or other medical disorder affecting the central nervous system. At each study timepoint, all participants with psychosis were administered the Positive and Negative Syndrome Scale (PANSS) [20] , Montgomery-Asberg Depression Rating Scale (MADRS) [21] , Young Mania Rating Scale (YMRS) [22], Clinical Anxiety Scale (CAS) [23], and the Brief Assessment of Cognition in Schizophrenia (BACS; tablet version [BAC] was used in the tES study) [24,25]. Detailed study procedures are outlined in Tamminga et al. [26].

### EEG procedures

EEG data were recorded using a 64-sensor cap with an extended 10/20 layout (Compumedics, Neuroscan), nose reference, and forehead ground. Sensor impedances were kept below 10kΩ and data were sampled at 1000Hz. A 5-minute eyes-open resting state was collected where participants were instructed to sit comfortably with their eyes loosely fixed on a white plus sign in the center of a screen.

### EEG data processing

Raw data were inspected for bad sensor recordings, interpolated (<10% of channels), transformed to an average reference, and digitally filtered from .5 Hz (12 dB/oct, zero phase) to 55 Hz (48 dB/oct, zero phase) with a notch filter at 60 Hz (2 Hz width) in BESA (MEGIS Software). Eye blinks, heart rate, and muscle tension artifacts were minimized using Independent Component Analysis in EEGLAB in MatLab (MathWorks). In EEGLAB, data were downsampled to 500 Hz and segmented in 5 second epochs. Epochs containing amplitudes +/-100µV at any sensor and datasets with fewer than 20 epochs were excluded from analysis.

Data were transformed into the time-frequency domain in EEGLAB by conducting Fast Fourier Transforms (FFTs) on Hanning tapered windows (500 ms steps, 1 Hz resolution) for each epoch. Power values (squared absolute values of complex FFT outputs) were then converted to decibels (10*log10) and averaged over time.

### EEG data reduction

To capture maximum explanatory variance across variables, avoid redundancy, and reduce statistical comparisons, frequency data were reduced by principal component analysis (PCA), as in our prior publication [5]. PCAs were conducted in the cross-sectional data and resulting weights were applied to all 3 studies. 55 frequencies were reduced to 4 bands: delta/theta (1-7Hz), alpha (8-15Hz), beta (16-30Hz), and gamma (31-55Hz). Spatial PCAs were performed on each frequency band to further reduce 64 sensor data to one virtual sensor. These PCAs were a near-identical replication of Thomas et al. [5], conducted in pseudo-resting state data, and are depicted in Figure 1. After PCA reduction, data were adjusted for age effects using the method by Dukart et al. [27]. In the cross-sectional healthy group only, quadratic and linear regressions were conducted on each PCA component. Components with significant age effects were adjusted for all participants and studies by subtracting the product of the regression coefficients and age for each individual.

**Figure 1.**
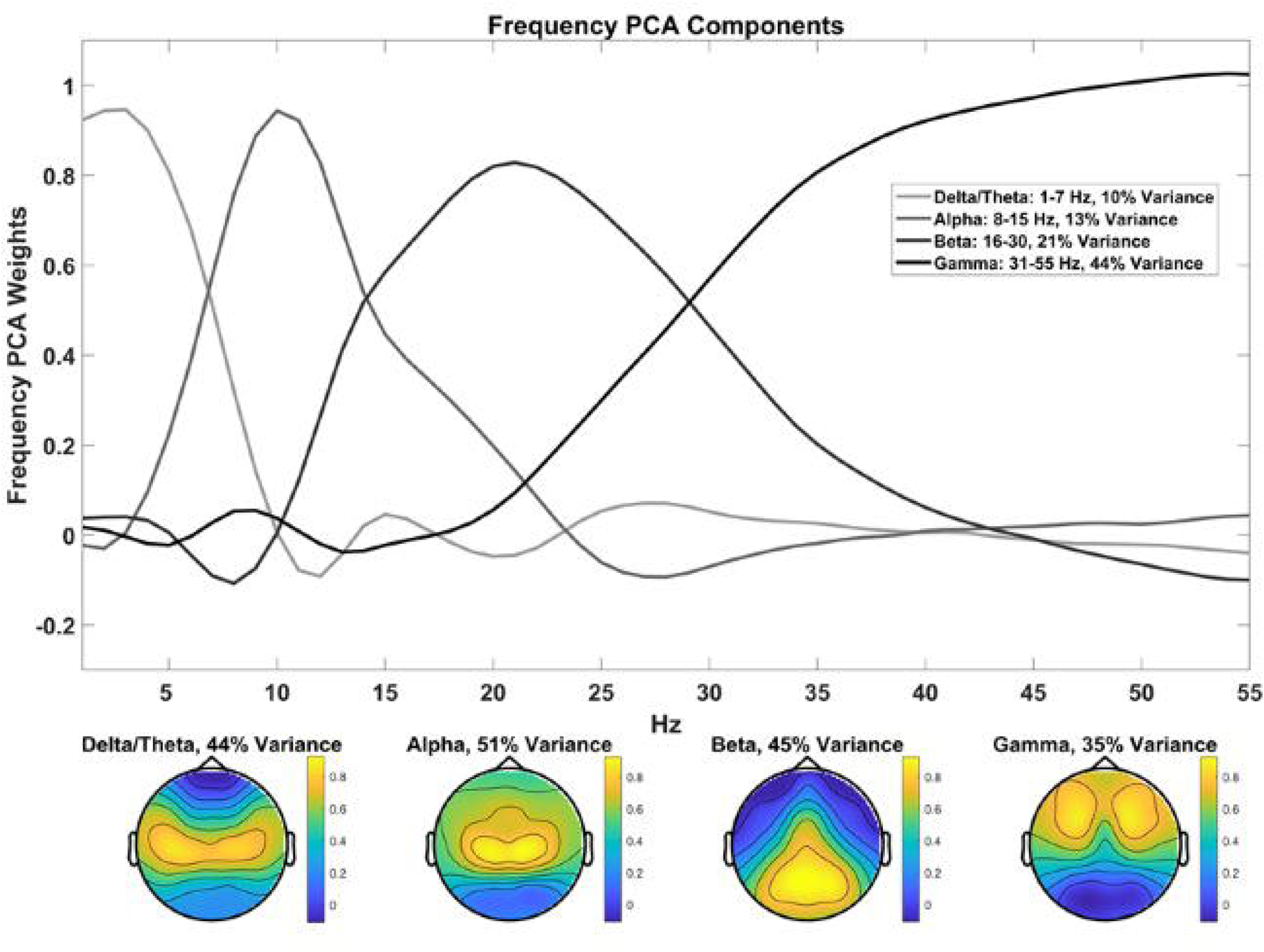
Frequency and spatial PCAs. Weights from the initial frequency PCA are shown at the top. Spatial PCAs were conducted for each frequency component and are shown at the bottom. PCAs reduced 55 frequencies and 64 sensors to one value per frequency band. **Alt text:** A line plot showing PCA weights at each frequency for the 4 frequency components and head topographies showing the distribution of PCA weights across the scalp for the 4 spatial components.

### tES target engagement procedure

This pilot intervention used an open-label crossover design with 2 HD-tES interventions: 1 targeting the left dlPFC (N=4) with an anode-center ring montage (anode F3 at 2mA, cathodes AF3, F5, F1, and FC3 at .50mA each) and 1 targeting the left TPJ (N=5) with a cathode-center ring montage (cathode P5 at 2mA, anodes C3, TP7, P1, and PO7 at .50mA each; Figure 4). All but 1 participant participated in both arms of the study (Table S3). For each arm, participants received 5 days of stimulation for 20 minutes twice daily, with a 2-week washout between arms. In each arm, EEG and clinical assessments occurred prior to the first stimulation session on day 1 and immediately following the final stimulation session on day 5. In addition to clinical assessments conducted in the longitudinal and cross-sectional studies, this study collected the Symptom Checklist-90 (SCL-90) [28] , a self-report instrument measuring domains of somatization, obsessive-compulsive, interpersonal sensitivity, depression, anxiety, hostility, phobic anxiety, paranoid ideation, and psychoticism. As this pilot study used an open-label design, participants, assessors, and tES administrators were not blind to study condition.

## Statistical Analyses

### Longitudinal stability statistics

Missing data was imputed using a random forest model in R Studio [29] (27% missing data, unimputed data reported in Table S5). Intraclass correlations (ICCs), which assess test-retest reliability [30], were conducted to test if frequency bands remain stable between baseline, 6-month, and 12-month timepoints. ICCs were conducted for all subjects and then separately for each group using 2-way mixed-effects models with absolute agreement. ICC values were compared between groups using Fisher’s z test to determine if stability differs between individuals with and without psychosis.

### Cross sectional statistics

Data were then entered into a mixed-design ANOVA with a 4-factor design: DSM X Biotype X sex X frequency, with alpha set at .05. Data violated Mauchly’s Test of Sphericity, so Greenhouse-Geisser corrections were applied. Follow-up ANOVAs and Tukey’s B tests were conducted as appropriate to identify homogenous subgroups while controlling for type-1 error, and Glass’ Delta effect sizes were calculated to quantify magnitude of effects. Finally, a bivariate Pearson correlation was conducted between average rsEEG power and the Intrinsic EEG Activity (IEA) biofactor reported in Parker et al. [10] to determine the degree of overlap between these two constructs.

### HD-tES target statistics

Wilcoxon signed-rank tests, the nonparametric equivalent of paired-samples t-tests, were conducted for each frequency band and symptom domain to test differences between pre- and post-stimulation timepoints. Alpha was set at .10 to reflect the exploratory, proof-of-concept aims typical of early target-engagement studies [31]. Within-subject effect sizes (ES, dz) were computed and power analyses were conducted in GPower to estimate required sample sizes for a future trial. Qualitatively, EEG difference topographies were examined to assess the spatial distribution of EEG changes.

## Results

### Longitudinal results

ICCs revealed good-to-excellent stability across bands and groups (ICC = .75–.95; Figure 3), with only four correlation coefficients falling below .80. Notably, the gamma frequency band was most stable for Biotype-1 (ICC = .88), the alpha and delta/theta frequency bands were most stable for Biotype-2 (ICC = .92), and the alpha frequency band was most stable for Biotype-3 (ICC = .95, Table S4). Results did not substantially differ for unimputed data (Table S5). In contrast, symptom measures were less stable over time, with moderate-to-good stability in PANSS and Social Functioning variables (ICCs = .64-.86) and poor-to-good stability in mood variables (ICCs = .34-.82; Figure 2).

**Figure 2.**
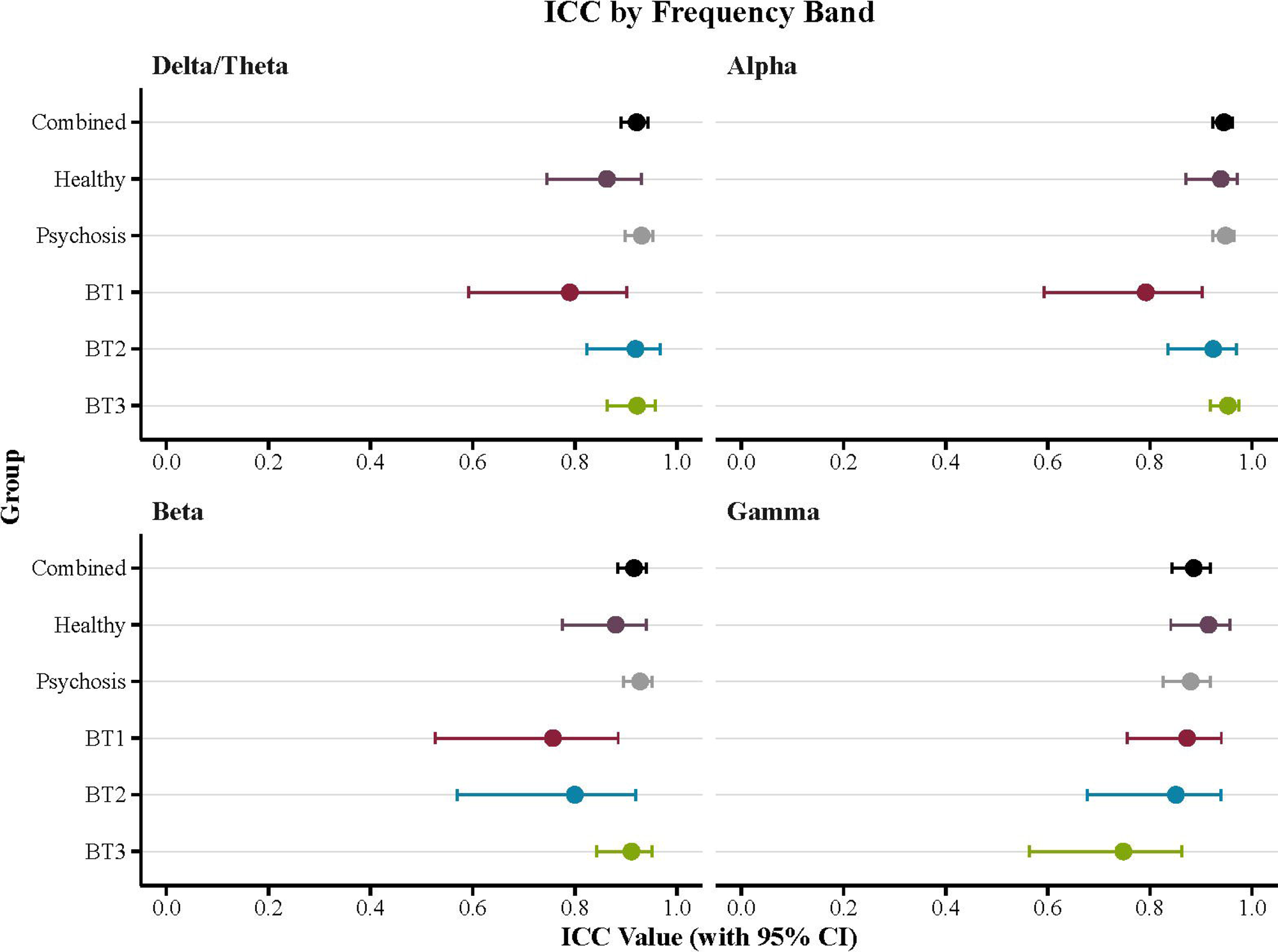
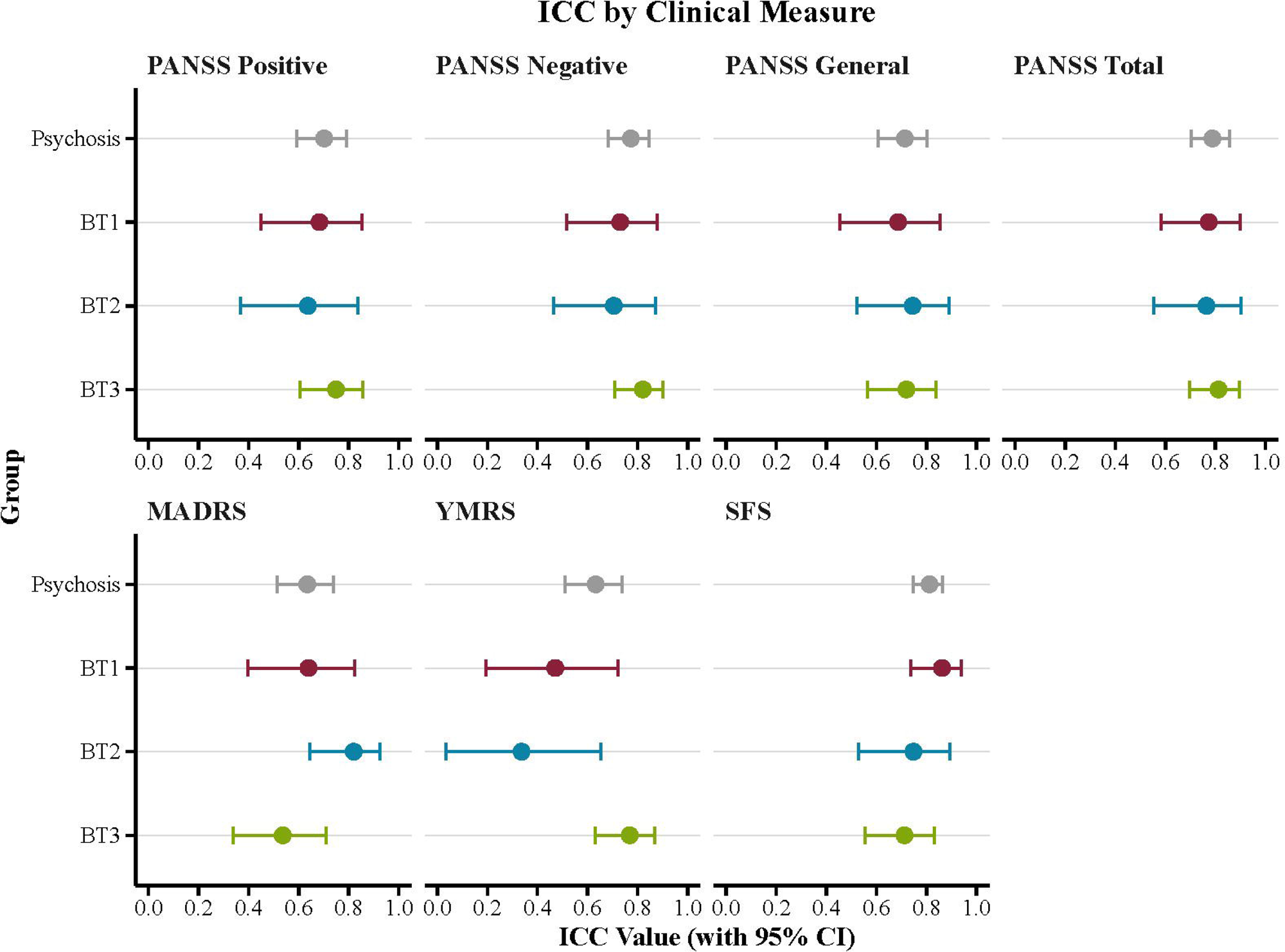
ICC values for rsEEG frequencies and symptoms by group. Dots signify the ICC value with a 95% confidence interval. rsEEG showed significant, good-to-excellent stability across all bands and groups. Sample sizes: Combined=109, Healthy=29, Psychosis=80, Biotype-1 (BT1)=25, Biotype-2 (BT2)=18, Biotype-3 (BT3)=37. **Alt text:** Scatter plots with 95% confidence interval error bars showing the estimated ICC for each group by frequency band and clinical measure.

**Figure 3.**
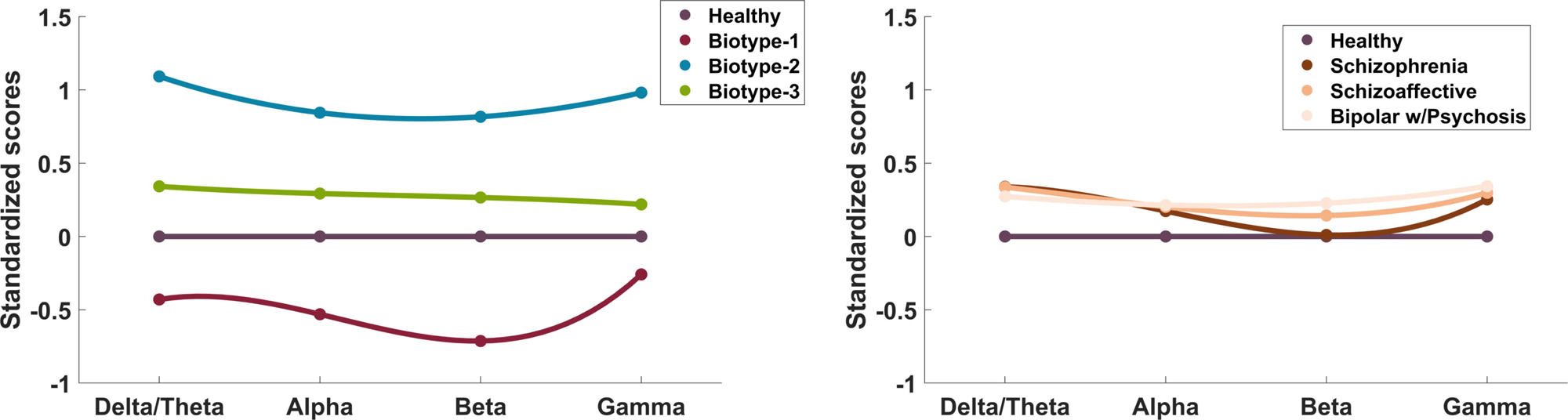
Glass Delta effect sizes of rsEEG for each group relative to healthy comparisons. Large effects are seen in Biotypes-1 and −2, with abnormally low activity in Biotype-1 and abnormally high activity in Biotype-2. rsEEG is mildly elevated in Biotype-3 and all DSM-categorized psychosis groups. **Alt text:** Line plots showing the effect size of each group at each frequency band organized by Biotype and DSM diagnosis.

**Figure 4.**
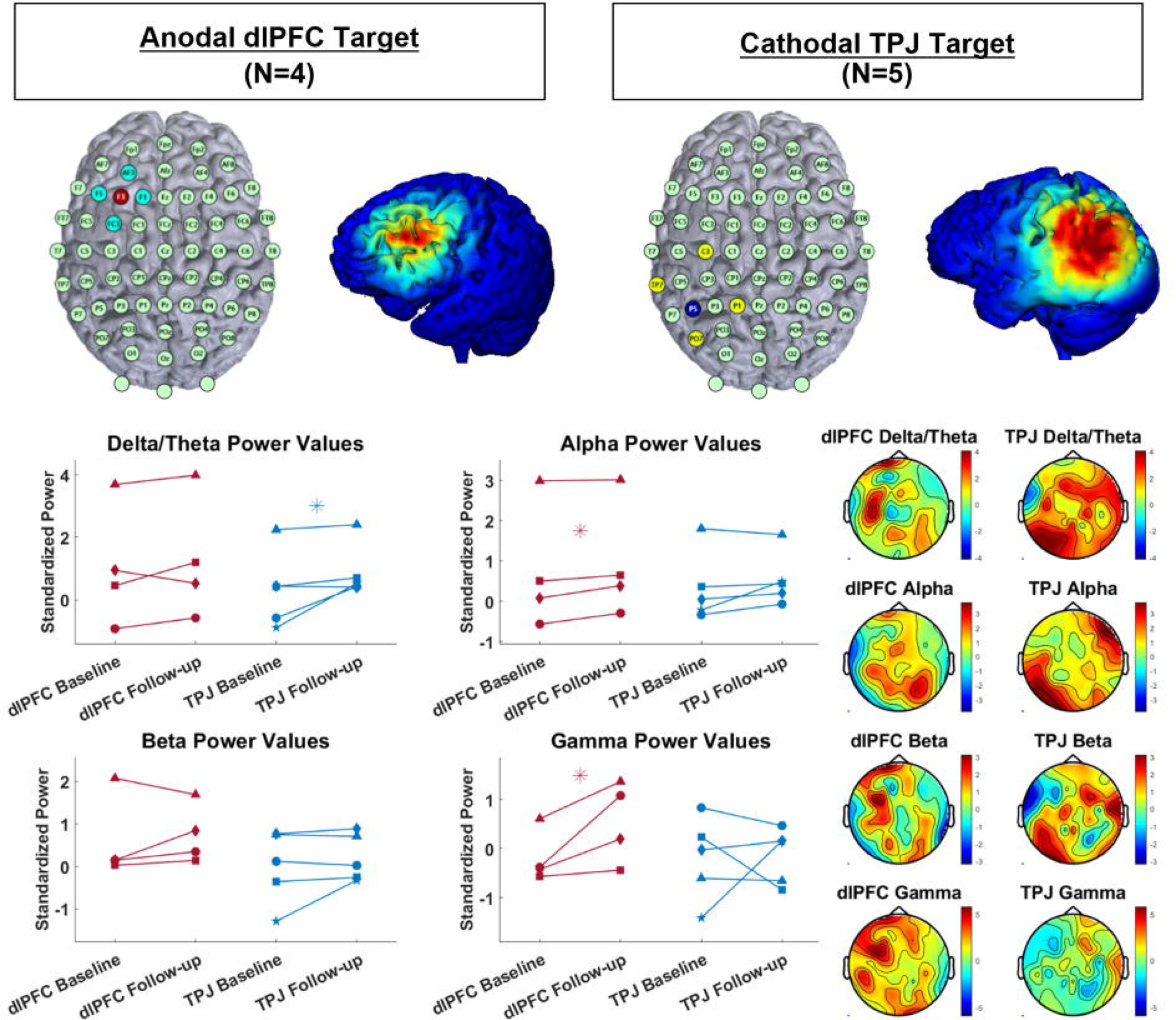
tES montages and rsEEG results. Montages (top) depict stimulating electrode locations and current flow models. Blue/teal electrodes are cathodes and red/yellow electrodes are anodes. Current flow models depict absolute values of injected current to illustrate focality. Results (bottom) show increased delta/theta power in the cathodal TPJ condition and increased alpha and gamma power in the anodal dlPFC condition (*p<.10). In topographies of the change values (follow-up minus baseline; right), enhanced power (red) is consistent with the site of stimulation. **Alt text:** A multi-paneled figure showing 1) a 64-sensor HD-tDCS cap layout highlighting the sensors used for each stimulation arm, 2) a brain depicting current flow on the cortex, 3) line plots showing each subject’s scores on outcomes for each condition, and 4) EEG topographies showing change in rsEEG power across scalp locations.

FDR corrected Fisher’s Z tests revealed that the gamma frequency band was less stable than the alpha frequency band in the total sample (*z* = –2.76, *p* = .03). Additionally, Biotype-1 was less stable than the healthy group in the alpha frequency band (*z* = –2.25, *p* = .012), and Biotype-3 was less stable than the healthy group in the gamma frequency band (*z* = –2.26, *p* = .012). No significant differences in stability were seen between the total sample of individuals with psychosis and the healthy group (all *p* > .05).

### Cross sectional results

In the cross-sectional sample, the ANOVA revealed a significant main effect of Biotype (F(2,2128) = 312.22, *p* < .001), where Biotype-1 < Healthy < Biotype-3 < Biotype-2. There was not a significant main effect of DSM (F(2,2128) = .89, *p* = .41) or between-subjects interactions (all *p* > .05). There was a significant frequency X Biotype interaction (F(3.96,4209.80) = 33.10, *p* < .001) such that beta oscillations were most impaired and gamma least impaired in Biotype-1 while delta/theta and gamma oscillations were most impaired in Biotype-2 (Figure 3). Full ANOVA results are reported in the supplement (Table S6). Finally, there was a significant correlation between average rsEEG power and the intrinsic activity biofactor reported in Parker et al. (r=.79, p<.001), indicating a high degree of overlap between these two constructs.

### HD-tES targeting results

In the dlPFC condition, alpha and gamma power increased after stimulation (both Z = - 1.83, *p* =.07, alpha ES = .79, gamma ES = 1.33). In the TPJ condition, delta/theta power increased (Z = 1.75, *p* = .08; Figure 4, ES = .90). Examination of change score topographies (follow-up – baseline) showed that changes primarily occurred around the stimulated region (left frontal and left posterior, respectively).

Behaviorally, participants in the dlPFC condition showed a decrease in PANSS Positive and Total scores (both Z = −1.84, *p* = .07, Positive ES = 2.51, Total ES = 2.02) and an increase in BAC Verbal Fluency (Z = 1.83, *p* = .07, ES = 1.93; Figure S1). In the TPJ condition, SCL-90 Psychoticism scores decreased (Z = −1.83, *p* = .07, ES = .94) and BAC Digit Sequencing scores increased (Z = −2.03, *p* = .04, ES = 1.24; Figure S1). No significant changes were observed in mood, anxiety, or other cognitive domains, and no significant side effects were reported (Figure S1).

A power analysis estimated how many participants are necessary for a future study to provide 80% power to 2-tailed Wilcoxon tests. For dlPFC stimulation, N = 7 per group is necessary to detect gamma change, N = 16 for the alpha change, and N = 4-5 for clinical and cognitive changes. For TPJ stimulation, N = 13 per group is necessary to detect the delta/theta change, N = 12 for the SCL-90 Psychoticism change, and N = 8 for the Digit Sequencing change.

## Discussion

We sought to test the stability, diagnostic specificity, and treatment relevance of rsEEG in psychotic disorders. In a longitudinal, non-intervention cohort, we showed that rsEEG is stable over 12 months. Then, in a large, cross-sectional cohort, we demonstrated that rsEEG substantially distinguishes Biotypes of psychosis from healthy controls and one another. Finally, in a pilot intervention, we demonstrated feasibility and preliminary proof-of-concept for targeting rsEEG with HD-tDCS in psychosis. Overall, these studies provide evidence that rsEEG is a clinically relevant biomarker with high potential for implementation in psychiatry.

Results from our longitudinal stability cohort demonstrated that rsEEG power values have good-to-excellent stability across 1-year. There were no significant differences in stability between healthy controls and people with psychosis. Additionally, rsEEG was more stable than symptom assessments, which showed poor-to-moderate test-retest-reliability, indicating that rsEEG remained stable even when symptoms fluctuated. Together, these findings indicate that rsEEG is more trait-like than state-like, reinforcing its suitability as a treatment related biomarker. It is possible that rsEEG power could display more changes over a longer course of time (for example, from early course to chronic illness) or with changes in medication, as some pharmaceuticals like Clozapine are associated with rsEEG changes. We are currently examining these hypotheses in ongoing studies.

The second study demonstrated that Biotype-1 displays abnormally low oscillatory power, Biotype-2 shows abnormally high power, and Biotype-3 has slightly high power. Effect sizes were large, with the most substantial effect on beta oscillations in Biotype-1 and on delta/theta oscillations in Biotype-2, though effects were largely consistent across all frequency bands. This suggests that differences are reflective of overall resting neural activity, rather than specific to any one frequency and its associated physiology. In contrast, there was not an effect of diagnosis (schizophrenia, bipolar disorder, schizoaffective disorder, healthy) on rsEEG power. This clarifies the mixed results and small effect sizes seen in previous literature [4], showing that rsEEG variance is not diagnosis-specific, but may be Biotype-specific. Results could point to differences in excitatory/inhibitory balance between Biotypes, as pilot data suggest glutamatergic hypoactivity in Biotype-1 and hyperactivity in Biotype-2 [32]. We will further examine these possibilities and other rsEEG metrics like microstates, graph metrics, or complexity in future studies.

Cross-sectional results replicate our group’s prior findings from rest-like EEG activity occurring prior to and after the primary evoked response to stimuli, termed “intrinsic” activity [5,9,11,33]. Specifically, we demonstrated a strong correlation (r = .79) between rsEEG and the intrinsic activity described in Parker et al. [10]. This is notable because rsEEG does not require the high temporal resolution equipment or software necessary for event-related EEG, making it a more practical biomarker to implement in a low-resourced or mobile environment, that still captures the same construct as intrinsic activity. Still, rsEEG alone is not adequate to make a Biotype diagnosis. Analysis is currently underway to develop an efficient algorithm for Biotype diagnosis [34,35]. These results suggest that rsEEG should be considered in such algorithms to streamline Biotype identification.

Finally, results from our pilot HD-tDCS study indicated that anodal stimulation of the left dlPFC may enhance alpha and gamma oscillations, and cathodal stimulation of the left TPJ may enhance delta/theta oscillations. Both stimulation approaches were associated with clinical improvements like reduced positive symptoms and enhanced verbal fluency following dlPFC stimulation and reduced psychoticism and enhanced digit sequencing following TPJ stimulation. rsEEG results of the dlPFC condition were consistent with our hypotheses, where anodal, excitatory-like current increased oscillatory power, however TPJ results did not follow the hypothesis of decreasing power, but rather increased low-frequency power. Recent evidence suggests that cathodal current may have a more complex effect on the neural membrane than simply hyperpolarization of cortex, as it may have some excitatory effects and modulate axonal fibers [36]. Given these considerations and the small sample size, more work is necessary to determine how tDCS affects resting oscillations. Results from the dlPFC arm support the initiation of a larger study examining the effects of HD-tDCS on rsEEG and symptoms in psychosis, particularly for individuals with low rsEEG power (Biotype-1).

The longitudinal and cross-sectional components of this study benefited from a large, transdiagnostic sample collected across differing geographic locations in the United States. The wide range of sociodemographic and clinical features captured by this sample supports the generalizability of findings, however findings may not generalize to other populations that are early-course, inpatient, unmedicated, or outside of the United States. The pilot tDCS component was intended as a preliminary feasibility study and thus had a small sample size without placebo control, constraining statistical power and risking expectancy/practice effects.

Additionally, the lack of counterbalancing study arms could mean that carryover effects of the first intervention (dlPFC) could have impacted results of the second (TPJ). Still, results support the initiation of a larger, placebo-controlled study to confirm and further investigate the potential efficacy of this therapy.

Overall, these investigations provide evidence that rsEEG is a strong, stable psychosis biomarker poised for clinical implementation. Easily collected rsEEG data may be highly beneficial for diagnosing psychosis Biotypes and used as a novel target for stratified neurophysiological interventions. Future research should focus on examining rsEEG fluctuations with pharmaceuticals, using rsEEG to stratify treatment approaches, and further investigating how tDCS may affect rsEEG oscillations and behavior.

## Disclosures

Authors Trotti, Doss, Parker, Raymond, Sauer, Hill, and Lizano report no financial relationships with commercial interests.

Drs. Pearlson, Gershon, Keedy, Tamminga, McDowell, Keshavan, and Clementz are on B-SNIP Diagnostics Board of Managers.

Dr. Gershon is a Consultant for Kynexis Corporation.

Dr. Tamminga is on the Kynexis Scientific Advisory Board and receives a retainer and the Karuna Therapeutics Scientific Advisory Board and owns stock.

Dr. Keshavan is an advisor for Alkermes.

Dr. Clementz is on the Kynexis Corporation Scientific Advisory Board.

## Data Availability

Cross-sectional and longitudinal data produced in the present work is available online at the NIMH Data Archive.
tDCS data produced in the present study is available upon reasonable request to the authors.

https://nda.nih.gov/edit_collection.html?id=2274

https://nda.nih.gov/edit_collection.html?id=2126

https://nda.nih.gov/edit_collection.html?id=2165

## Acknowledgments

B-SNIP2: R01MH077851, R01MH103368, R01MH077945, R01MH103366, R01MH078113 PARDIP: R01MH096900, R01MH096942, R01MH096957, R01MH096913

B-SNIP1: R01MH078113, R01MH077945, R01MH077862, R01MH077851, R01MH077852 UL1TR002378, TL1TR002382, F32MH135669

## References

[1] Deco G, Jirsa VK, McIntosh AR. Emerging concepts for the dynamical organization of resting-state activity in the brain. Nat Rev Neurosci 2011;12:43–56. 10.1038/nrn2961.

[2] Winterer G, Ziller M, Dorn H, Frick K, Mulert C, Wuebben Y, et al. Schizophrenia: reduced signal-to-noise ratio and impaired phase-locking during information processing. Clinical Neurophysiology 2000;111:837–49.

[3] Rolls ET, Loh M, Deco G, Winterer G. Computational models of schizophrenia and dopamine modulation in the prefrontal cortex. Nat Rev Neurosci 2008;9:696–709. 10.1038/nrn2462.

[4] Newson JJ, Thiagarajan TC. EEG Frequency Bands in Psychiatric Disorders: A Review of Resting State Studies. Front Hum Neurosci 2019;12. 10.3389/fnhum.2018.00521.

[5] Thomas O, Parker DA, Trotti RL, McDowell JE, Gershon ES, Sweeney JA, et al. Intrinsic neural activity differences in psychosis biotypes: Findings from the Bipolar-Schizophrenia Network on Intermediate Phenotypes (B-SNIP) consortium. Biomark Neuropsychiatry 2019;1:100002. 10.1016/j.bionps.2019.100002.

[6] Narayanan B, Soh P, Calhoun VD, Ruaño G, Kocherla M, Windemuth A, et al. Multivariate genetic determinants of EEG oscillations in schizophrenia and psychotic bipolar disorder from the BSNIP study. Transl Psychiatry 2015;5. 10.1038/tp.2015.76.

[7] Raymond N, Lizano P, Kelly S, Hegde R, Keedy S, Pearlson GD, et al. What can clozapine’s effect on neural oscillations tell us about its therapeutic effects? A scoping review and synthesis. Biomark Neuropsychiatry 2022;6. 10.1016/j.bionps.2022.100048.

[8] Popov T, Tröndle M, Baranczuk-Turska Z, Pfeiffer C, Haufe S, Langer N. Test–retest reliability of resting-state EEG in young and older adults. Psychophysiology 2023;60. 10.1111/psyp.14268.

[9] Clementz BA, Parker DA, Trotti RL, McDowell JE, Keedy SK, Keshavan MS, et al. Psychosis Biotypes: Replication and Validation from the B-SNIP Consortium. Schizophr Bull 2022;48:56–68. 10.1093/schbul/sbab090.

[10] Parker DA, Trotti RL, McDowell JE, Keedy SK, Keshavan MS, Pearlson GD, et al. Differentiating biomarker features and familial characteristics of B-SNIP psychosis Biotypes. Transl Psychiatry 2025;15:281.

[11] Hudgens-Haney ME, Ethridge LE, Knight JB, McDowell JE, Keedy SK, Pearlson GD, et al. Intrinsic neural activity differences among psychotic illnesses. Psychophysiology 2017;54:1223–38. 10.1111/psyp.12875.

[12] Cheng PWC, Louie LLC, Wong YL, Wong SMC, Leung WY, Michael AN, et al. The effects of transcranial direct current stimulation (tDCS) on clinical symptoms in schizophrenia: A systematic review and meta-analysis. Asian J Psychiatr 2020;53. 10.1016/j.ajp.2020.102392.

[13] Brunelin J, Adam O, Mondino M. Recent advances in noninvasive brain stimulation for schizophrenia. Curr Opin Psychiatry 2022;35:338–44. 10.1097/YCO.0000000000000809.

[14] Boudewyn MA, Scangos K, Ranganath C, Carter CS. Using prefrontal transcranial direct current stimulation (tDCS) to enhance proactive cognitive control in schizophrenia. Neuropsychopharmacology 2020;45:1877–83. 10.1038/s41386-020-0750-8.

[15] Kostova R, Cecere R, Thut G, Uhlhaas PJ. Targeting cognition in schizophrenia through transcranial direct current stimulation: A systematic review and perspective. Schizophr Res 2020;220:300–10. 10.1016/j.schres.2020.03.002.

[16] Adam O, Blay M, Brunoni AR, Chang HA, Gomes JS, Javitt DC, et al. Efficacy of Transcranial Direct Current Stimulation to Improve Insight in Patients With Schizophrenia: A Systematic Review and Meta-analysis of Randomized Controlled Trials. Schizophr Bull 2022;48:1284–94. 10.1093/schbul/sbac078.

[17] Gainsford K, Fitzgibbon B, Fitzgerald PB, Hoy KE. Transforming treatments for schizophrenia: Virtual reality, brain stimulation and social cognition. Psychiatry Res 2020;288. 10.1016/j.psychres.2020.112974.

[18] Parlikar R, Chhabra H, Selvaraj S, Shivakumar V, Sreeraj VS, Dinakaran D, et al. Neurobiological and clinical effects of High-Definition tDCS on persistent auditory hallucinations in schizophrenia: A randomized controlled trial. MedRxiv [Preprint] 2023. 10.1101/2023.05.10.23289796.

[19] First MB, Spitzer RL, Gibbon M, Williams JB. Structured Clinical Interview for DSM-IV-TR Axis I Disorders, Research Version, Patient Edition (SCID-I/P) 2002.

[20] Opler LA, Kay SR, Lindenmayer JP, Fiszbein A. Structured Clinical Interview: The Positive and Negative Syndrome Scale (SCI-PANSS) 1999.

[21] Williams JBW, Kobak KA. Development and reliability of a structured interview guide for the Montgomery-Åsberg Depression Rating Scale (SIGMA). British Journal of Psychiatry 2008;192:52–8. 10.1192/bjp.bp.106.032532.

[22] Young RC, Biggs JT, Ziegler VE, Meyer DA. A rating scale for mania: Reliability, validity and sensitivity. British Journal of Psychiatry 1978;133:429–35.

[23] Snaith R, Baugh S, Clayden A, Hussain A, Sipple M. The Clinical Anxiety Scale: A modification of the Hamilton Anxiety Scale. British Journal of Psychiatry 1982;141:518–23. 10.1192/bjp.141.5.518.

[24] Keefe RSE, Harvey PD, Goldberg TE, Gold JM, Walker TM, Kennel C, et al. Norms and standardization of the Brief Assessment of Cognition in Schizophrenia (BACS). Schizophr Res 2008;102:108–15. 10.1016/j.schres.2008.03.024.

[25] Keefe RSE, Goldberg TE, Harvey PD, Gold JM, Poe MP, Coughenour L. The Brief Assessment of Cognition in Schizophrenia: Reliability, sensitivity, and comparison with a standard neurocognitive battery. Schizophr Res 2004;68:283–97. 10.1016/j.schres.2003.09.011.

[26] Tamminga CA, Ivleva EI, Keshavan MS, Pearlson GD, Clementz BA, Witte B, et al. Clinical phenotypes of psychosis in the Bipolar-Schizophrenia Network on Intermediate Phenotypes (B-SNIP). American Journal of Psychiatry 2013;170:1263–74. 10.1176/appi.ajp.2013.12101339.

[27] Dukart J, Schroeter ML, Mueller K, The Alzheimer’s Disease Neuroimaging Initiative. Age correction in dementia - Matching to a healthy brain. PLoS One 2011;6:1–9. 10.1371/journal.pone.0022193.

[28] Derogatis LR. The SCL-90-R. Baltimore: Clinical Psychometric Research 1975. 10.1002/9780470479216.corpsy0970.

[29] Buuren S van, Groothuis-Oudshoorn K. mice: Multivariate Imputation by Chained Equations in R. J Stat Softw 2011;45:1–67. 10.18637/jss.v045.i03.

[30] Koo TK, Li MY. A Guideline of Selecting and Reporting Intraclass Correlation Coefficients for Reliability Research. J Chiropr Med 2016;15:155–63. 10.1016/j.jcm.2016.02.012.

[31] Lee EC, Whitehead AL, Jacques RM, Julious SA. The statistical interpretation of pilot trials: Should significance thresholds be reconsidered? BMC Med Res Methodol 2014;14. 10.1186/1471-2288-14-41.

[32] Bolo NR, Zeng V, Parker D, Ivleva E, McDowell J, Keedy S, et al. Hippocampal glutamate and verbal episodic memory in the psychosis spectrum: A preliminary report. Biomark Neuropsychiatry 2025;13. 10.1016/j.bionps.2025.100135.

[33] Clementz BA, Sweeney JA, Hamm JP, Ivleva EI, Ethridge LE, Pearlson GD, et al. Identification of distinct psychosis biotypes using brain-based biomarkers. American Journal of Psychiatry 2016;173:373–84. 10.1176/appi.ajp.2015.14091200.

[34] Clementz BA, Chattopadhyay I, Hill SK, McDowell JE, Keedy SK, Parker DA, et al. Cognitive performance and differentiation of B-SNIP psychosis Biotypes: Algorithmic Diagnostics for Efficient Prescription of Treatments (ADEPT) - 2. Biomark Neuropsychiatry 2025;12. 10.1016/j.bionps.2024.100117.

[35] Clementz BA, Chattopadhyay I, Trotti RL, Parker DA, Gershon ES, Hill SK, et al. Clinical characterization and differentiation of B-SNIP psychosis Biotypes: Algorithmic Diagnostics for Efficient Prescription of Treatments (ADEPT)-1. Schizophr Res 2023;260:143–51. 10.1016/j.schres.2023.08.006.

[36] Qi S, Cao L, Wang Q, Sheng Y, Yu J, Liang Z. The Physiological Mechanisms of Transcranial Direct Current Stimulation to Enhance Motor Performance: A Narrative Review. Biology (Basel) 2024;13. 10.3390/biology13100790.

