## Supplement for "Evaluating Resting State EEG Biomarkers Across Psychosis Biotypes: Stability and HD-tDCS Modulation"

Supplementary Information

| **Table S1.** Clinical Characteristics of the Cross-Sectional Sample. | | | | |
| --- | --- | --- | --- | --- |
| **Characteristic** | **Biotype-1**  **N = 476*^1^*** | **Biotype-2**  **N = 447*^1^*** | **Biotype-3**  **N = 478*^1^*** | **p-value*^2^*** |
| **Positive and Negative Syndrome Scale (PANSS)** | 63 (19) | 67 (20) | 57 (19) | <0.001 |
| PANSS Positive | 16 (6) | 18 (6) | 14 (6) | <0.001 |
| PANSS Negative | 15 (6) | 16 (7) | 13 (6) | <0.001 |
| PANSS General | 31 (9) | 33 (10) | 30 (9) | <0.001 |
| **Montgomery-Asberg Depression Scale** | 12 (10) | 11 (10) | 11 (10) | >0.9 |
| **Young Mania Rating Scale** | 9 (8) | 9 (8) | 8 (7) | 0.002 |
| **Birchwood Social Functioning Scale (SFS)** | 121 (24) | 118 (23) | 129 (24) | <0.001 |
| **BACS** | -1.80 (1.22) | -2.17 (1.23) | -0.62 (1.14) | <0.001 |
| *^1^*Mean (SD) | | | | |
| *^2^*Kruskal-Wallis rank sum test | | | | |

| **Table S2.** Additional Clinical Characteristics of the Cross-Sectional Sample. | | | | |
| --- | --- | --- | --- | --- |
| **Characteristic** | **Biotype-1 N = 476** | **Biotype-2 N = 447** | **Biotype-3 N = 478** | **p-value*^1^*** |
| **Mean Illness Duration in Years (SD)** | 19 (13) | 20 (12) | 17 (12) | 0.003 |
| **Medication Status (%)** |  |  |  | 0.5 |
| Not Medicated | 5.4% | 4.3% | 6.0% |  |
| Medicated | 95% | 96% | 94% |  |
| **Mean Medication Count (SD)** | 4.8 (3.4) | 5.0 (3.5) | 4.6 (3.2) | 0.2 |
| **Mean Psychotropic Medication Count (SD)** | 2.84 (1.61) | 2.92 (1.53) | 2.85 (1.54) | 0.5 |
| **Psychotropics (%)** |  |  |  | 0.080 |
| Off | 7.1% | 7.5% | 11% |  |
| On | 93% | 92% | 89% |  |
| **Antipsychotics (%)** |  |  |  | <0.001 |
| Off | 16% | 16% | 28% |  |
| On | 84% | 84% | 72% |  |
| **1st Generation Antipsychotics (%)** |  |  |  | 0.010 |
| Off | 88% | 86% | 92% |  |
| On | 12% | 14% | 7.9% |  |
| **2nd Generation Antipsychotics (%)** |  |  |  | 0.004 |
| Off | 24% | 26% | 33% |  |
| On | 76% | 74% | 67% |  |
| **Antidepressants (%)** |  |  |  | >0.9 |
| Off | 54% | 54% | 54% |  |
| On | 46% | 46% | 46% |  |
| **Tricyclics (%)** |  |  |  | >0.9 |
| Off | 98% | 98% | 98% |  |
| On | 1.5% | 1.8% | 1.7% |  |
| **SSRIs (%)** |  |  |  | 0.7 |
| Off | 69% | 67% | 69% |  |
| On | 31% | 33% | 31% |  |
| **Other Antidepressants (%)** |  |  |  | 0.8 |
| Off | 78% | 80% | 79% |  |
| On | 22% | 20% | 21% |  |
| **Mood Stabilizers (%)** |  |  |  | 0.004 |
| Off | 57% | 58% | 48% |  |
| On | 43% | 42% | 52% |  |
| **Lithium (%)** |  |  |  | 0.063 |
| Off | 89% | 84% | 84% |  |
| On | 11% | 16% | 16% |  |
| **Anticonvulsants (%)** |  |  |  | 0.009 |
| Off | 65% | 70% | 60% |  |
| On | 35% | 30% | 40% |  |
| **Anxiolytics/Sedatives/Hypnotics(%)** |  |  |  | 0.5 |
| Off | 77% | 77% | 80% |  |
| On | 23% | 23% | 20% |  |
| **Anticholinergics (%)** |  |  |  | <0.001 |
| Off | 87% | 82% | 92% |  |
| On | 13% | 18% | 8.3% |  |
| **Stimulants (%)** |  |  |  | 0.072 |
| Off | 95% | 95% | 92% |  |
| On | 4.7% | 4.6% | 7.7% |  |
| **Mean CPZ Equivalent Dose (SD)** | 548 (855) | 523 (683) | 452 (754) | 0.067 |
| *^1^*Kruskal-Wallis rank sum test; Pearson's Chi-squared test | | | | |

| **Table S3.** tES Sample Demographics. | | | | | |
| --- | --- | --- | --- | --- | --- |
| Subject | Biotype | Diagnosis | Age | Sex | Baseline GAF |
| 1 | Biotype-1 | Schizoaffective | 38 | Male | 75 |
| 2*^1^* | Biotype-1 | Schizophrenia | 22 | Male | 75 |
| 3 | Biotype-3 | Bipolar w/Psychosis | 31 | Female | 71 |
| 4 | Biotype-3 | Schizophrenia | 54 | Male | 65 |
| 5 | Biotype-3 | Schizophrenia | 27 | Female | 55 |
| *^1^*TPJ condition only | | | | | |

| **Table S4.** ICC with 95% CI by Group and Frequency Band | | | | | | | | |
| --- | --- | --- | --- | --- | --- | --- | --- | --- |
|  |  |  | 95% CI | | F Test with True Value 0 | | | |
| Group | Frequency Band | *ICC* | *LB* | *UB* | *F* | *df1* | *df2* | *p* |
| Combined | Delta/Theta | .92 | .89 | .94 | 12.77 | 108 | 216 | <.001 |
| Healthy | Delta/Theta | .86 | .75 | .93 | 7.88 | 28 | 56 | <.001 |
| Psychosis | Delta/Theta | .93 | .90 | .95 | 14.35 | 79 | 158 | <.001 |
| Biotype-1 | Delta/Theta | .79 | .59 | .90 | 4.7 | 24 | 48 | <.001 |
| Biotype-2 | Delta/Theta | .92 | .82 | .97 | 12 | 17 | 34 | <.001 |
| Biotype-3 | Delta/Theta | .92 | .86 | .96 | 13.87 | 36 | 72 | <.001 |
| Combined | Alpha | .95 | .92 | .96 | 18.67 | 108 | 216 | <.001 |
| Healthy | Alpha | .94 | .87 | .97 | 20.73 | 28 | 56 | <.001 |
| Psychosis | Alpha | .95 | .92 | .97 | 19.11 | 79 | 158 | <.001 |
| Biotype-1 | Alpha | .79 | .59 | .90 | 4.67 | 24 | 48 | <.001 |
| Biotype-2 | Alpha | .92 | .84 | .97 | 13.47 | 17 | 34 | <.001 |
| Biotype-3 | Alpha | .95 | .92 | .97 | 22.92 | 36 | 72 | <.001 |
| Combined | Beta | .92 | .88 | .94 | 12.18 | 108 | 216 | <.001 |
| Healthy | Beta | .88 | .78 | .94 | 9.14 | 28 | 56 | <.001 |
| Psychosis | Beta | .93 | .90 | .95 | 13.97 | 79 | 158 | <.001 |
| Biotype-1 | Beta | .76 | .53 | .89 | 4.04 | 24 | 48 | <.001 |
| Biotype-2 | Beta | .80 | .57 | .92 | 5.44 | 17 | 34 | <.001 |
| Biotype-3 | Beta | .91 | .84 | .95 | 12.37 | 36 | 72 | <.001 |
| Combined | Gamma | .89 | .84 | .92 | 8.73 | 108 | 216 | <.001 |
| Healthy | Gamma | .92 | .84 | .96 | 12.76 | 28 | 56 | <.001 |
| Psychosis | Gamma | .88 | .83 | .92 | 8.25 | 79 | 158 | <.001 |
| Biotype-1 | Gamma | .87 | .76 | .94 | 7.82 | 24 | 48 | <.001 |
| Biotype-2 | Gamma | .85 | .68 | .94 | 6.87 | 17 | 34 | <.001 |
| Biotype-3 | Gamma | .75 | .56 | .86 | 3.9 | 36 | 72 | <.001 |

| **Table S5.** ICC with Unimputed Data | | | | | | | | |
| --- | --- | --- | --- | --- | --- | --- | --- | --- |
|  |  |  | 95% CI | | F Test with True Value 0 | | | |
| Group | Frequency Band | *ICC* | *LB* | *UB* | *F* | *df1* | *df2* | *p* |
| Combined | Delta/Theta | 0.95 | 0.92 | 0.96 | 18.74 | 76 | 154 | <.001 |
| Healthy | Delta/Theta | 0.94 | 0.87 | 0.98 | 17.54 | 16 | 33 | <.001 |
| Psychosis | Delta/Theta | 0.95 | 0.92 | 0.97 | 19.06 | 59 | 120 | <.001 |
| Biotype-1 | Delta/Theta | 0.85 | 0.67 | 0.94 | 6.30 | 18 | 37 | <.001 |
| Biotype-2 | Delta/Theta | 0.93 | 0.82 | 0.97 | 13.28 | 13 | 27 | <.001 |
| Biotype-3 | Delta/Theta | 0.95 | 0.91 | 0.98 | 24.20 | 26 | 42 | <.001 |
| Combined | Alpha | 0.96 | 0.94 | 0.97 | 23.54 | 76 | 152 | <.001 |
| Healthy | Alpha | 0.97 | 0.92 | 0.99 | 34.59 | 16 | 24 | <.001 |
| Psychosis | Alpha | 0.95 | 0.93 | 0.97 | 22.29 | 59 | 119 | <.001 |
| Biotype-1 | Alpha | 0.85 | 0.68 | 0.94 | 6.52 | 18 | 37 | <.001 |
| Biotype-2 | Alpha | 0.95 | 0.89 | 0.98 | 21.61 | 13 | 28 | <.001 |
| Biotype-3 | Alpha | 0.96 | 0.92 | 0.98 | 25.15 | 26 | 51 | <.001 |
| Combined | Beta | 0.94 | 0.92 | 0.96 | 17.43 | 76 | 153 | <.001 |
| Healthy | Beta | 0.96 | 0.90 | 0.98 | 29.18 | 16 | 25 | <.001 |
| Psychosis | Beta | 0.94 | 0.90 | 0.96 | 16.08 | 59 | 117 | <.001 |
| Biotype-1 | Beta | 0.83 | 0.64 | 0.93 | 5.90 | 18 | 38 | <.001 |
| Biotype-2 | Beta | 0.82 | 0.55 | 0.94 | 6.43 | 13 | 21 | <.001 |
| Biotype-3 | Beta | 0.93 | 0.87 | 0.97 | 14.98 | 26 | 49 | <.001 |
| Combined | Gamma | 0.89 | 0.84 | 0.93 | 8.86 | 76 | 153 | <.001 |
| Healthy | Gamma | 0.95 | 0.88 | 0.98 | 23.86 | 16 | 23 | <.001 |
| Psychosis | Gamma | 0.87 | 0.81 | 0.92 | 7.83 | 59 | 119 | <.001 |
| Biotype-1 | Gamma | 0.87 | 0.72 | 0.94 | 7.46 | 18 | 37 | <.001 |
| Biotype-2 | Gamma | 0.85 | 0.64 | 0.95 | 6.69 | 13 | 28 | <.001 |
| Biotype-3 | Gamma | 0.77 | 0.56 | 0.89 | 4.28 | 26 | 54 | <.001 |

| **Table S6.** Full ANOVA Results of Cross-Sectional Analysis. | | | |
| --- | --- | --- | --- |
| **Source** | **df** | **F** | **Sig.** |
| **Between-Subjects Effects** |  |  |  |
| Biotype | 2.00 | 306.58 | 0.00*** |
| DSM | 2.00 | 0.87 | 0.42 |
| Sex | 1.00 | 25.20 | 0.00*** |
| Biotype * DSM | 4.00 | 1.09 | 0.36 |
| Sex * Biotype | 2.00 | 1.82 | 0.16 |
| Sex * DSM | 2.00 | 0.14 | 0.87 |
| Sex * Biotype * DSM | 4.00 | 1.44 | 0.22 |
| Error | 2211.00 |  |  |
| **Within-Subjects Effects** |  |  |  |
| Frequency | 1.99 | 4913.66 | 0.00*** |
| Frequency * Biotype | 3.98 | 33.16 | 0.00*** |
| Frequency * DSM | 3.98 | 1.53 | 0.19 |
| Frequency * Sex | 1.99 | 5.12 | 0.01** |
| Frequency * Biotype * DSM | 7.96 | 0.70 | 0.69 |
| Frequency * Sex * Biotype | 3.98 | 1.24 | 0.29 |
| Frequency * Sex * DSM | 3.98 | 0.55 | 0.70 |
| Frequency * Sex * Biotype * DSM | 7.96 | 1.19 | 0.30 |
| Error(Frequency) | 4397.59 |  |  |

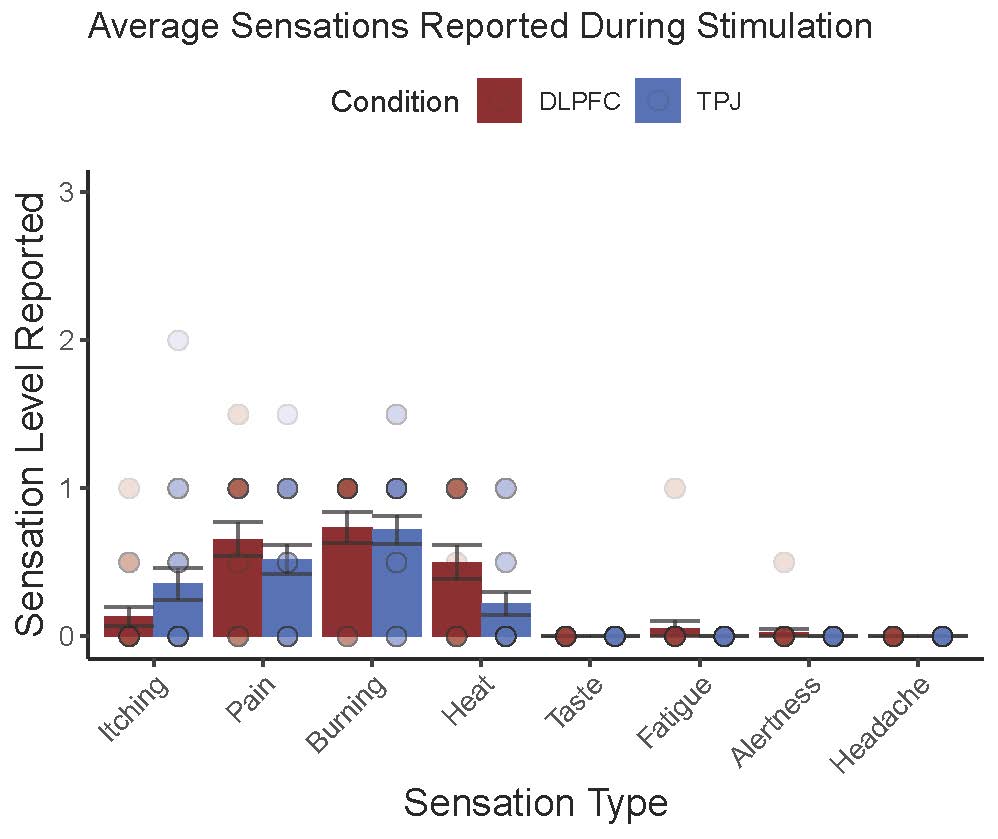

**Figure S1.** Data collected from side effect questionnaire after each session of HD-tDCS. Level 1=Mild, 2=Moderate, 3=Severe. There were no significant differences in sensations between conditions (all p>.30)
